# University of Texas at Arlington (UTA) in-house COVID-19 testing program

**DOI:** 10.1101/2021.09.08.21263264

**Authors:** Roy Rudewick, Taylor Gunby, Anajane G. Smith, Zibiao Guo, Angela J. Middleton, Necole Cox, Florence P. Haseltine

**Author notes:** Please send correspondence to: Florence P Haseltine, PhD, MD, Presidential Distinguished Professor of Nursing, Medical Director, North Texas Genome Center, University of Texas at Arlington, Arlington, Texas 76019.

## Abstract

The pandemic caused by the spread of the virus SARS-CoV-2 threatened to severely disrupt the activities of student-athletes. (REF) In order to provide a safe environment for athletic competition, the National Collegiate Athletic Association (NCAA) mandated testing of student-athletes. The goal was to rapidly identify student-athletes and the athletic staff member who either tested positive for SARS-CoV-2 or were in contact with individuals who tested positive. Rapid identification of infected individuals and their contacts allowed the University to implement quarantine standards and quarantine facilities quickly as needed. The University of Texas at Arlington (UTA) developed an in-house testing program and was quickly able to meet the NCAA requirements, allowing UTA to continue its athletic participation with minimal forfeiture of scheduled games. The purpose of this paper is to report the implementation UTA’s COVID prevention program for the university’s athletic program. This program will provide valuable information to other universities’ planning for the management of COVID prevention in their athletic programs. Challenges and solutions are identified.

## Introduction

During the 2020-2021 academic year, our programs and particularly the intercollegiate athletic programs faced a hurdle because of the COVID-19 pandemic (1,2). The programs had to maintain a safe environment for the student-athletes and athletic department staff in which to work and to compete. Student-athletes were at higher risk of exposure to the SARs-CoV-2 virus by nature of their activities and, potentially, had more opportunities to spread the virus if infected. The health of the student-athletes and athletic department staff was a major concern as more and more information on the effects of COVID-19 infection emerged (3). Our experience has influenced the development of our plan to keep our athletes and athletic program staff protected from COVID while returning to a fully open campus with a full athletic schedule in the upcoming academic year. This paper shares UTA’s experience to help inform other athletic departments and universities plans.

At the University of Texas at Arlington, our goal was to develop a rapid testing program allowing the student-athletes to be tested and results reported back within 24 to 48 hours in order to reduce COVID exposure and transmission. UTA is a member of the NCAA Sun Belt Division I. The NCAA provided guidelines (1) and rules for testing student-athletes to ensure that exposure to the SARS-CoV-2 virus was minimized for all concerned. Both molecular rtPCR tests and antigen tests for the virus could be used. The testing requirements that were established required 3 tests for SARS-CoV-2 each week, of which at least 2 were rtPCR tests, while one could be an antigen test. Delivering the testing results quickly was critical so that the teams could practice, travel, compete without interruption, or, when necessary, isolate.

### Description of Program

To facilitate the testing and reporting, UTA decided that testing would be done primarily site, an option that was available because the campus’s North Texas Genome Center (NTGC) was certified to perform human clinical diagnostic testing. This certification was fortuitously granted in January 2020, after application to and approval by two regulatory entities known as CLIA (4) and CAP (College of American Pathologists) (5).

Shortly after UT Arlington closed most of the campus to onsite instruction in March 2020, the administration began implementing the on-site COVID-19 testing program for the UTA community by acquiring of the necessary materials and equipment. One company, the Quidel Corporation, was beneficial by giving the NTGC access to supplies and reagents on the day after their Lyra-Direct SARS-CoV-2 Assay (6) received FDA Emergency Use Authorization. The Quidel Corporation provided the testing reagents and sample collection materials reliably throughout the entire testing period.

## Methods

From March through June 2020, the laboratory became proficient with the rtPCR test technology for SARs-CoV-2 analysis. Nasopharyngeal swab collections were performed at the University’s Athletic Center by trained staff. The samples were sent to the laboratory no later than 5 hours after sampling. The specimens were stored at 4°C in NTGC refrigerators before analysis. All the tests at the NTGC used the molecular rtPCR test Lyra-Direct SARS-CoV-2 Assay (Quidel, Inc). During the holidays, tests were done either by a third-party provider or with FDA EUA compliant COVID-19 rapid antigen tests to ensure the required COVID-19 testing standards were met for the student-athletes to continue to compete. A total of 10110 clinical tests were performed of which approximately 8000 for 388 students-athletes and 150 staff and volunteers associated with the athletic programs. The number of tests per individual was extremely variable. Some had only 1 and one individual had 36. The people who tested positive were not retested for over three months, and that was often after the requirements for their sport was over. Those under surveillance but never tested positive were continuously tested throughout the competitive season.

Screening, primarily of staff, started in June 2020, extending to the athletes in July of 2020. (Figure 1) The timing enabled the intercollegiate team sports to be tested routinely and return the results to the Senior Associate Athletics Director for Sports Medicine; greater than 29% (2974) of all tests were reported the same day, and more than 61% (6123) were completed and reported within 24 hours. In no case were results delayed more than 48 hours. Other UTA activity groups, such as the SPIRIT, the Movin Mav’s (wheelchair basketball) teams, ROTC and Band also participated in the program. Other students who remained on campus were also tested; many could not leave the area and attended hybrid classes when they were offered. This rapid reporting enabled the efficient identification of infected students.

**Figure 1:**
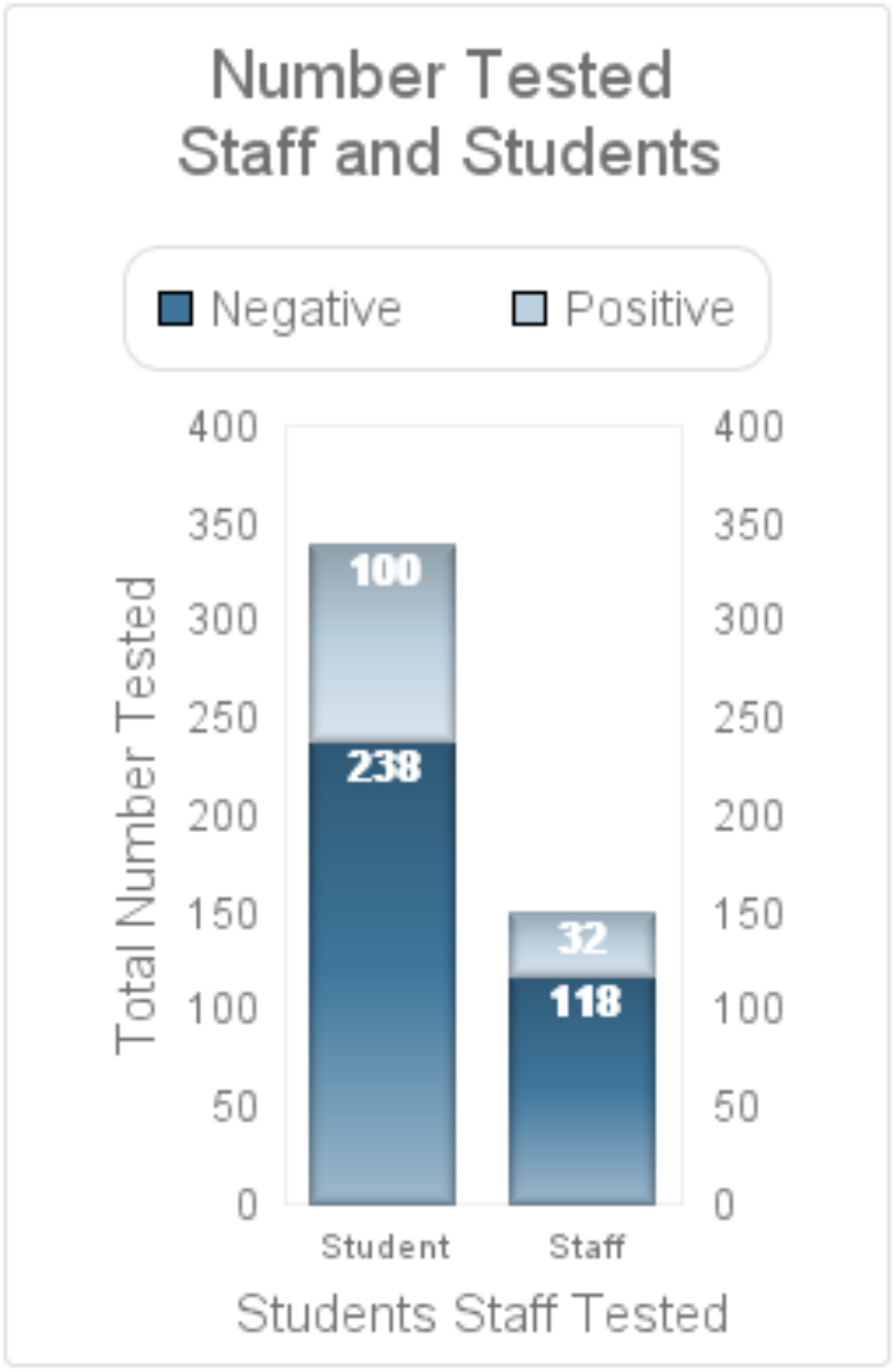
Shows the number of individual students and staff that were tested. 338 students were tested frequently. The number of tests per person ranged from 1 test (those individuals were positive and were quarantined, to 36 times for individuals who needed continuous surveillance.) 238 student-athletes remained negative throughout the testing period and 100 student-athletes tested positive for SARS-CoV-2. All of the 150 athletic staff (composed of faculty, trainers, coaches, volunteers, and student trainees) were tested at least once. 32 tested positive at some point for SARS-CoV-2.

UTA students who tested positive by NTGC were reported to the athletic department and student health services. The positive and negative tests were reported both to Tarrant County and to the Texas Department of Health (7). Working together, the UTA College of Nursing and Health Innovation and College of Social Work, with the assistance of the Student Medical Service, provided a robust contact tracing system and a method for housing and feeding the students who tested positive as well and their contacts. Individuals who were members of the UT Arlington staff were given their results directly by the NTGC and those individuals and their results were also conveyed to the County and State. The student-athletes and contacts were notified by the Student Health Services. NCAA requires a degree of medical transparency for competitive sports. Students and staff sign a release allowing their medical conditions to be shared with athletic programs. The staff with a positive result for COVID were individually notified by the NTGC and referred to their physician.

## Results

Providing our student-athletes with rapid testing and reporting enabled them to continue competing with only one missed game. 29% of our tests were reported the same day as testing, and 61% were reported by the following day. The remaining tests 10% were reported on day 2. We found that over the year almost 1/3 of our tested athletes did test positive for SARS-CoV-2. Intensive contact tracing and isolating of their contacts, 2/3 of the people appeared not to have contacted COVID during the testing period. As a result of rapid reporting, enabling isolation of infected individuals and exposed contacts all the teams were able to complete their competitive season. A total of 6 games had to be rescheduled as a result of UTA COVID positive student-athletes. UTA missed only on track meet. Other competitions that were rescheduled or cancelled were the result of COVID infections from other competitors. The athletic program accounted for 8,000 out of the 10110 tests run. (See distribution of results by activity in Figure 2)

**Figure 2:**
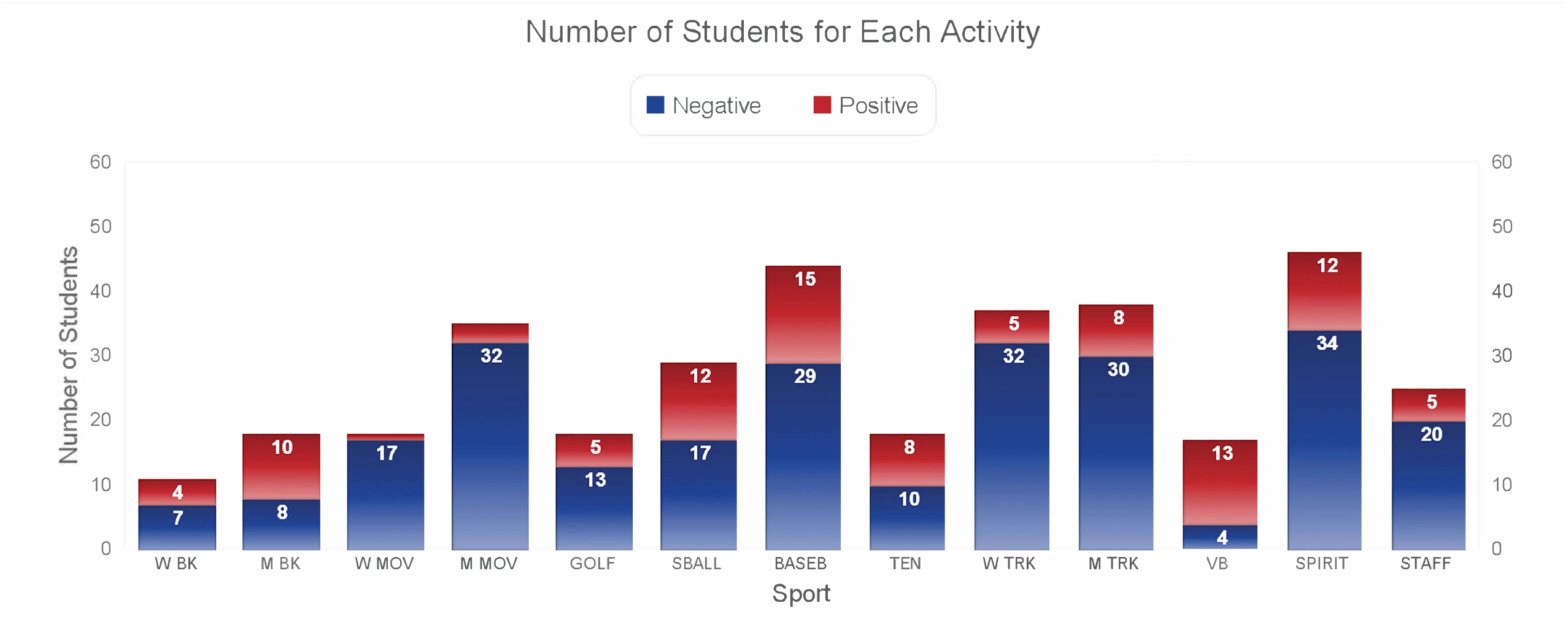
Shows the number of students tested and the results in for each athletic activity. The captions are W BK- Women’s Basketball, M BK- Men’s Basketball, W Mov – Women’s Movin Mavs, M Mov- Men’s Movin Mavs, Golf – Men’s & Women’s Golf, SBall - Softball, BaseB –Baseball, Ten – Men’s & Women’s Tennis, W Trk – Women’s Track, M Trk – Men’s Track,VB- Women’s Volleyball, Spirit- Spirit Teams, Staff- Student Staff

## Discussion

The ability to perform testing at the University initially offered two significant benefits. The first was rapid testing and reporting of the tests results enabling the University to immediately isolate and quarantine the infected individuals, as well as their exposed contacts. The second was cost. The University provided the resources to enhance the NTGC. Still much of the equipment was already present and the staff was very eager to contribute by running the university testing program. The logistics required a university-wide effort. The administration coordinated the laboratory testing needs by involving multiple UTA campus resources, including purchasing, the Office of Information Technology, Student Health Services, and others.

During the year there were a variety of challenges, such as complications due to holidays as well as changes in testing requirements. When the laboratory could not do the testing, as for example, during the Winter break, the athletic teams program made other arrangements.

## Conclusion

The SARS-CoV-2 testing program at the NTGC succeeded in providing timely tests for the athletes. The number of student-athletes that tested positive was highest in September, December, and January. (Figure 3) Although1 scheduled event had to be forfeited, UTA was able to field teams to meet its scheduled event. An additional unanticipated benefit was the ability of the student-athletes to attend hybrid classes in person and continue their education uninterrupted. This was also a positive development for faculty members who were giving lectures because it added to the number of those taking classes in person.

**Figure 3:**
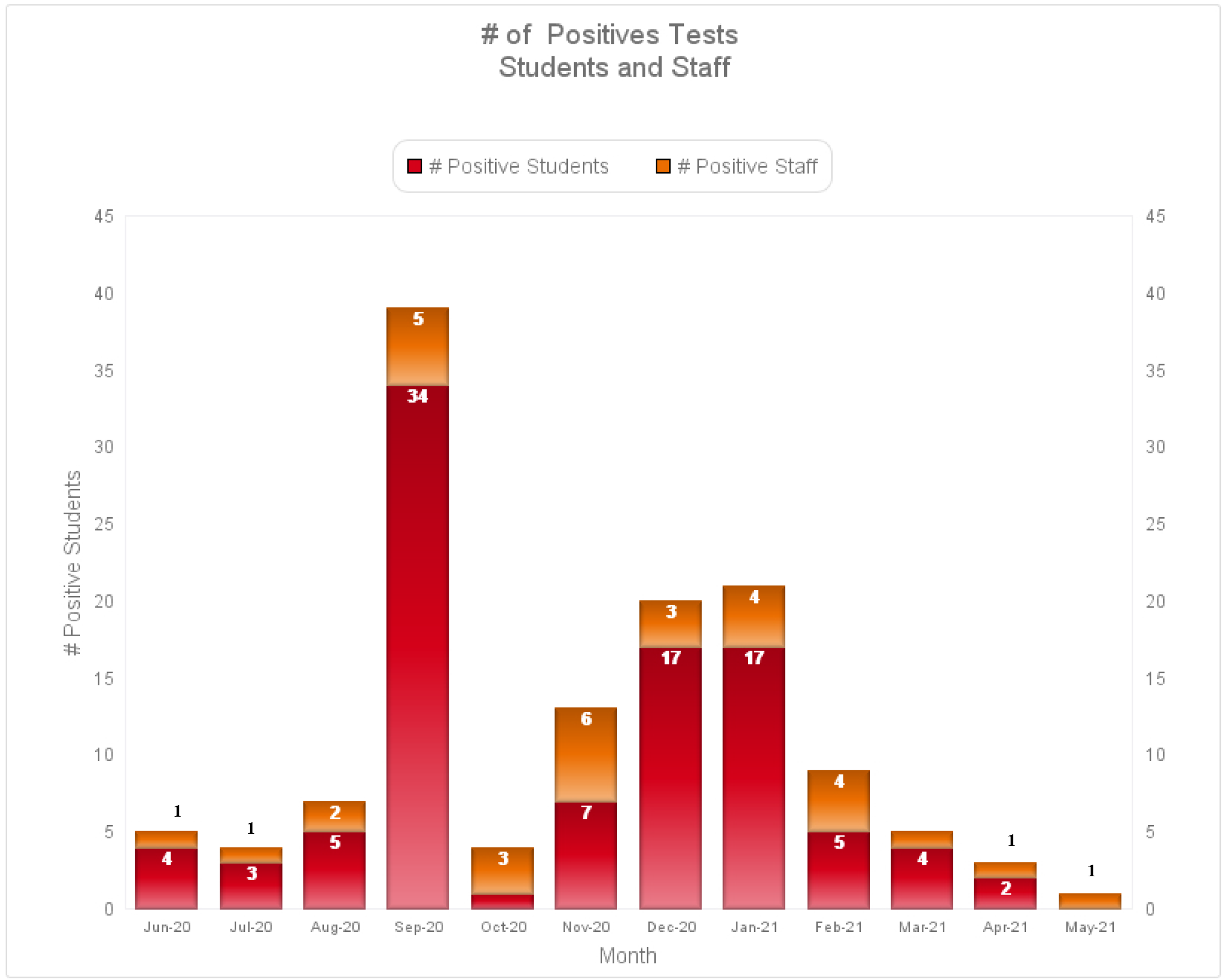
Indicates the number of negatives and positives for the students and staff by month tested.

## Data Availability

There was no research performed. Simple reporting of results.

## Acknowledgments

The results of the study are presented clearly, honestly, and without fabrication, falsification, or inappropriate data manipulation and do not constitute endorsement by the American College of Sports Medicine. Funds were provided by the University of Texas at Arlington. Dr. James Grover and the other senior administrations really pushed this through. The help of Edward Gonzales in the Office of Information Technology at UTA was outstanding.

## Notes

### Competing Interest Statement

The authors have declared no competing interest.

### Clinical Trial

This is a report of routine test results. They were not part of a trial.The testswere mandated for student athletes by the National Collegiate Athletic Association (NCAA) in order for the students to participate in sports.

### Funding Statement

Institutional funds from the University of Texas Arlington were used to support this work
All investigators were supported by the UTA

### Author Declarations

The Institutional Review Board at the University of Texas at Arlington. Consent was obtain at the time the students signed up for intercollegiate sports activities

